# Note: Forecasting COVID-19 spread in Lebanon February 8-21,2021

**DOI:** 10.1101/2021.02.09.21251447

**Authors:** Omar El Deeb, Maya Jalloul

## Abstract

This note implements an iterative method in order to predict the number of active cases with COVID-19, and consequently forecast the number of inpatients to hospitals and ICU in Lebanon according to different scenarios after end of the complete closure and curfew implemented between January 13 and February 7, 2021. The forecast predicts a decrease in the number of infections and people in need for hospitalization and ICU during the 2 weeks after the curfew (until February 21st), with varying extents depending on the subsequent commitment to mitigation measures, except for the case of a 2% absolute increase from the current rate of infection, which would bring back the numbers of cases back to a increasing trend.

## 1 Introduction

The evolution spead of COVID19 in Lebanon has experienced several stages and key dates. Approximately one year after recording the first case on February 21, 2020, the cumulative number of cases has reached 319917, including 3616 deaths by February 7, 2021 [1,2]. The peak number of daily infections was recorded on January 15 at 6154 new cases, which exceeds the total number of infections recorded in the first five and a half months of the disease spread in Lebanon. This was the result of a sharp surge in the rate of infections during the end of December. Soon after, the government imposed a countrywide strict lockdown starting on January 13 until February 7, followed by four phases of gradual lifting of measures [3].

The sharp rise in infections was accompanied by a sharp rise in the hospitalized patients as well as those in the intensive care units (ICU). Despite the expansion of the COVID19 units as well as the increase in the number of beds that have been implemented over the year, first in the public health sector, then in the private hospitals, officials have repeatedly declared that hospitals reached their saturation levels [4],[3].

In this context, this note aims to examine the potential changes expected in the rates of infection, hospitalization, ICU occupancy and use of respirators, in light of the first phase of lifting the measures. The methodolgy involves the use an iterative model that relies on the infection data of the two weeks prior to a specific point in time, February 8 in this case, in order to forecast the short-term changes up to two following weeks, as in [5,6]. We investigate four diferent scenarios which refiect diferent levels of committment to social distancing measures. Our results suggest that high commitment would substantially decrease the pressure on the health care sector, while a lower degree committment would restore the surge in infections as well as hospitalizations in a significant short period of time, reversing the results of the strict lockdown.

## 2 The iterative method

The iterative method uses the available daily data of infections and recoveries to forecast progression of the disease. The active infected daily cases are denoted by *I*_*i*_ where *i* is the index of days and *i* ∈ [1, *n*]. We use the last *m* values of *I*_*i*_ to determine the average arithmetic gross rate in the last *m* days as

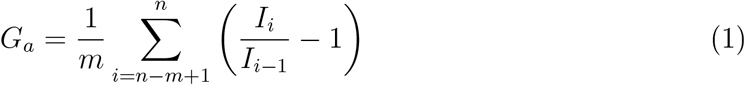

and the average geometric gross rate of the same set of data as

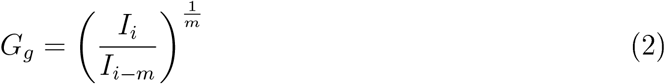

*G*_*a*_ and *G*_*g*_ indicate the average rate of increase in the number of current infections during the last *m* days, using arithmetic and geometric means. We forecast the progression of the number of infections by *I*_*i*+1_ = *I*_*i*_ (1 + *G*_*a*_) using the arithmetic gross infection rate *G*_*a*_ or alternatively *I*_*i*+1_ = *I*_*i*_*G*_*g*_ using the geometric gross infection rate *G*_*g*_.

To account for the removal of active cases, we define the death and recovery rates by *p* and 1 − *p* respectively, the number of days need for recovery by *h* and the average number of days between infection and death by *d*. This implies that on day (*i* + 1), the number of deaths will be *p* (*I*_*i*−*d*_ − *I*_*i*−*d*−1_) where (*I*_*i*−*d*_ − *I*_*i*−*d*−1_) represents the number of people who caught the virus *d* days ago. Similarly, the number of people recovered would be proportional to the number of people who caught the virus *h* days ago and is given by (1 − *p*) (*I*_*i*−*h*_ − *I*_*i*−*h*−1_). The values of *p* and *h* vary in the literature and in the available data from the country under consideration. Here we take *p* = 0.02, *h* = 20 days and *d* = 24 days.

**Figure 1:**
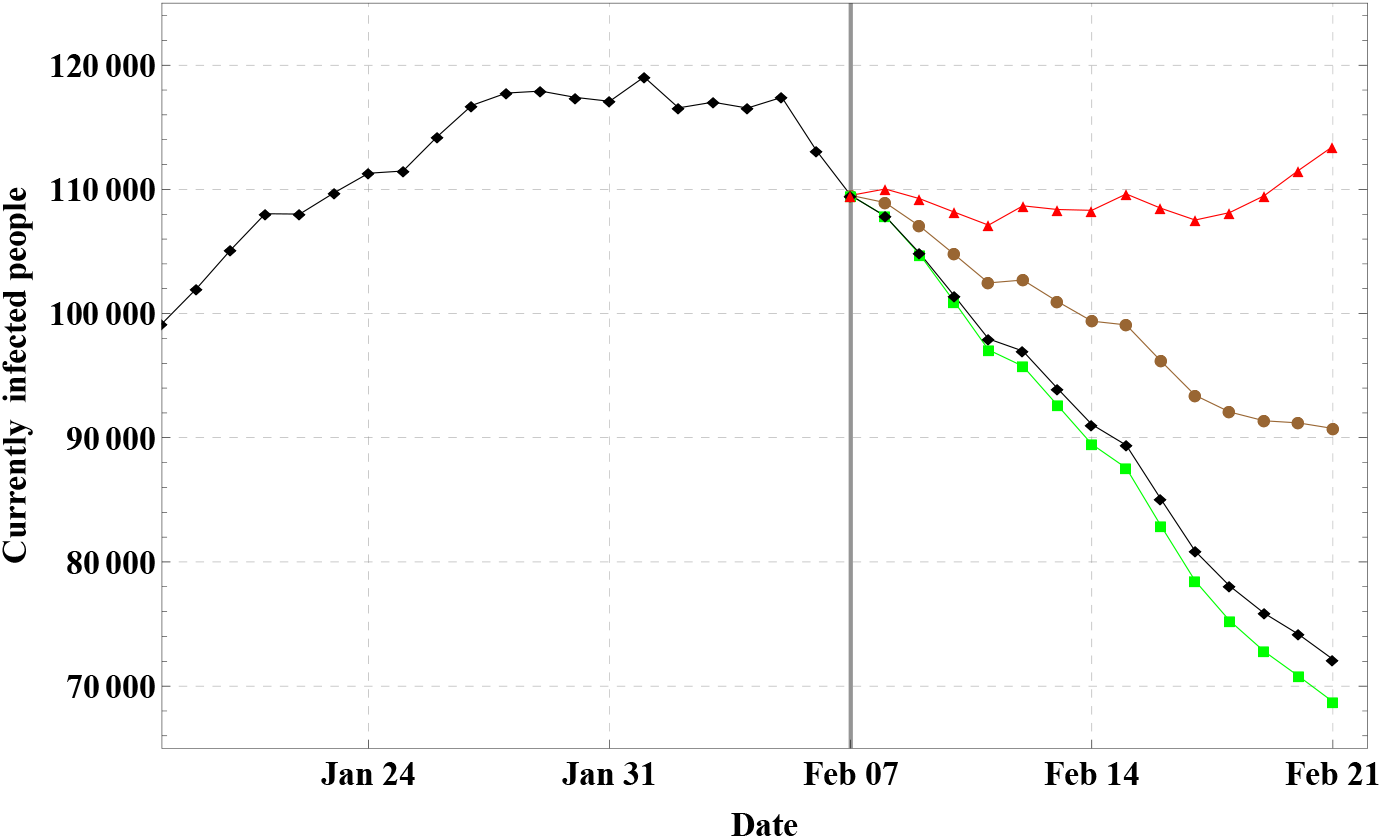
A plot of the four possible simulated scenarios for the number of active cases between February 7 and February 21. The green and balck lines simulate possible cases if the lockdown would continue fully applied while the brown and red lines represent respectively the number of active cases as expected due to an absolute 1% and 2% increase in the infecttion rates.

The recursive relation is defined as follows:

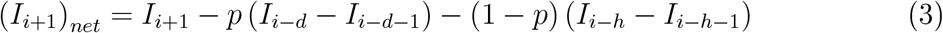

where (*I*_*i*+1_)_*net*_is the net number of infected people on day (*i* + 1). In every iteration, we use the available data up to the day under consideration, estimate the number of infections on the next day, and reevaluate *G*_*a*_ and *G*_*g*_ accordingly, hence recursively predicting the progression of the rate and the number of active infected cases. We perform our simulations using *G*_*a*_, *G*_*g*_ and possible increases of these rates.

## 3 Results and conclusion

The results revealed from the simulations of the four forecasts (See 1) between February 7 and 21, 2021 show that the continuation of spread rates similar to the last 14 days precedding February 7 would lead to a substantial decline in the number of active cases, while a 1% aboslute increase in the rates would still keep a relative decline in the number of active cases until February 21, despite a possible rise afterwards. Contrarily, less mitigation measures leading to an absolute increase of 2% to the current infection rates would soon bring back the level of acive cases close to their levels in the first week of February.

**Figure 2:**
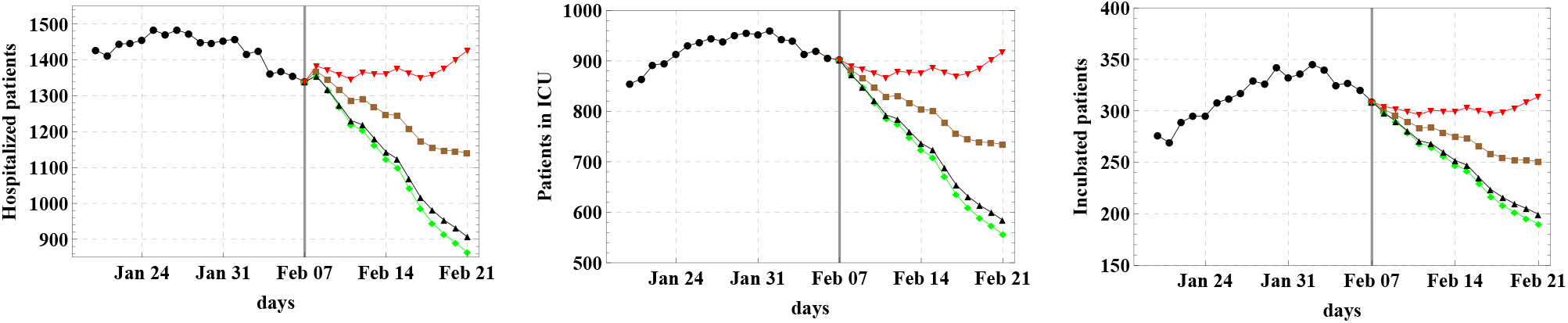
The 3 adjacent figures represent the numbers of expected hospitalized cases (left), ICU cases (middle) and intubated cases (right) according to four possible scenarios where The green and balck lines in each figure forecast the number of hospitalized, ICU or intubated cases if the lockdown would continue fully applied while the brown and red lines represent the forecast of those three cases as as expected due to an absolute 1% and 2% increase in the infecttion rates.

The number of hospitalized patients, ICU patients and intubated patients follow a similar general trend as shown in. The number of hospitalized, ICU or intubated cases would substantially decrease if the lockdown would continue fully applied while the decrease is relatively less under the assumption of a 1% absolute increase in infection rates, and a quick return to the high levels recorded during the first week of February is predicted if the infection rates increase by a rate of 2%.

The number of hospitalized, ICU and intubated patients is closely related to the number of active cases as statistics on recorded data reveal. This fact is exploited, together with the iterative method used in the simulation of active cases, in order to forecast the number of the three levels of the needed patient care.

Previous forecasts on the number of cases in Lebanon has proven to be successful and the actual data were in range of expectations according to the models implemented [7]. This supports the motivation behind current forecasts in order to supply the decision makers with some estimates that are necessary for policy measures to be undertaken.

## Data Availability

Data available upon request.

## Notes

### Competing Interest Statement

The authors have declared no competing interest.

### Funding Statement

The authors have no funding.

### Author Declarations

Public data used.

